# Computed Tomography-Based Analysis of Foramen Magnum and Occipital Condyles in adult Nigerians: An Anatomical Study

**DOI:** 10.1101/2023.07.15.23292716

**Authors:** Fatima Muhammad, Umar Ahmad, Tawfiq Yousef Tawfiq ZYOUD, Murtala Muhammad Jibril, Suleiman Hamidu Kwairanga

## Abstract

**Background:** Precise identification of craniometric points is pivotal in forensic and anthropological research for reliable and accurate gender determination. The foramen magnum and occipital condyles’ morphology offers valuable data for this purpose.

**Aim:** The study aimed to determine variations in the morphology of the foramen magnum and occipital condyles among a population of 100 individuals, using advanced Computed Tomography (CT) scan technology.

**Methods:** We assessed a prospective cohort of 100 individuals who underwent cranial CT scans. The study focused on the morphological variations of the foramen magnum and occipital condyles, recording shapes such as oval, round, egg, tetragon, pentagon, hexagon, and irregular for the foramen magnum, and kidney-type, oval, S-like, 8-like, ring-like, and triangular for the occipital condyles.

**Results:** Notably, males demonstrated higher measurement values for most parameters, with the exception of the width of the occipital condyle, where females presented larger dimensions. Furthermore, a strong positive correlation was observed between the measurements of the foramen magnum (length and width) and specific parameters of the occipital condyles.

**Conclusions:** The study underscores the utility of foramen magnum dimensions, especially length, as a determinant in gender identification, with a success rate of 68% in the studied population. This research enhances our understanding of cranial morphology and its applications in forensic and anthropological investigations.

## Introduction

Foramen magnum (FM) is the largest foramen of the skull located in an anteromedian position which consequently leads into the posterior cranial fossa. It has the largest diameter termed as anteroposterior diameter; it is usually oval in shape with a broad base. It comprises the distal end of the medulla oblongata, meninges, vertebral arteries, and the spinal accessory nerve (Standring *et al*., 2008; Gautam *et al*., 2012; Muralidhar *et al.,* 2014; Babu, 2016). The foramen magnum shapes differs. The shape variations is of great importance as it has effect on the vital structures that passes through it due to the role it plays in different surgical approaches. Dimensions of the foramen magnum also have clinical importance because the vital structures that pass through it may suffer compression; therefore, the cranial base is such a difficult structure that is only studied morphometrically (Murshed *et al.,* 2003; Suazo *et al.,* 2009; Kumar, 2015; Babu, 2016).

In conditions of natural or man-made disasters where direct identification of victims is difficult, there is a need for an accurate, timely and simple method for gender determination (Teixeira *et al.,* 1982; Vinutha et al., 2018; TY *et al.,* 2020). The anatomical characteristics of the foramen magnum and occipital condyle also differ in race, regions within the same race and may also varied in periods of evolution. Moreover, the anatomical characteristics of the FM and occipital condyle have been found to vary across races, regions, and even evolutionary periods, thus reflecting the complexity and diversity of these structures (Catalina, 1987; Murshed *et al*., 2003; Singh *et al*., 2017).

Despite the potential importance of these variations, data is limited concerning the morphological characterization of the FM and occipital condyle among specific ethnic groups, such as those residing in the Bauchi metropolis. FM assessments bear significant implications not only for tailoring suitable surgical procedures but also for forensic medicine, where they contribute to identifying unknown gender and potentially ascertaining past trauma or causes of death (Erdil, *et al*., 2010; Fatma *et al*., 2010; Edward *et al*., 2013; Salih *et al*., 2015; El-Barrany *et al*., 2016).

Thus, this study aims to analyse the FM and occipital condyles measurements as potential indicators for skull gender identification, with a focus on the Nigerian population of the Bauchi metropolis. This research not only addresses the paucity of data on this topic, but it also endeavours to provide valuable insights to clinicians, researchers, and forensic scientists, ultimately enhancing our understanding of these critical anatomical landmarks.

## Materials and Methods

### Ethical clearance

Ethical clearance for this prospective study was granted by two authoritative bodies: the Bauchi State University Ethics Committee (BASUG/FBMS/REC/VOL. 2/005) and the Bauchi State Specialist Hospital’s Research and Ethical Committee (NREC/0/11/19B/2021/23). Data were accordingly collected from the Radiology Department of the Specialist Hospital in Bauchi, Nigeria, adhering to all ethical guidelines and practices mandated by these committees.

### Selection criteria

### Inclusion criteria

The study included individuals who met the following criteria:

1. Possession of cranial Computed Tomography (CT) scans.
2. No recorded history of cranial trauma.
3. Presentation of an intact foramen magnum and occipital condyles in the CT scans.
4. Clear visibility of the bony margins of the foramen magnum and occipital condyles in the CT scans.

### Exclusion criteria

The study excluded individuals based on the following criteria:

1. Presence of a disrupted or damaged foramen magnum and occipital condyles in cranial Computed Tomography (CT) scans.
2. Evidence of diseases leading to cranial deformities.
3. History of accidents, surgery, or pathologies involving the foramen magnum region.
4. Any disorder potentially impacting the normal anatomy of the skull base.

Only individuals in good health, having completed skeletal growth in adulthood, were considered for inclusion in the study.

### Sampling frame

The study’s sampling frame consisted of Computed Tomography (CT) scan samples procured from the Radiology Department of the Specialist Hospital in Bauchi, Nigeria. The acquisition of these samples was facilitated by obtaining written permission from the relevant authorities before the commencement of the data collection process.

### Sample size and study subjects

This prospective study utilized cranial Computed Tomography (CT) scans from a random selection of individuals aged between 21 and 65 years, retrieved from the Radiology Department of Specialist Hospital Bauchi. Sample size was determined based on the formula by Lwanga and Lemeshaw (1991), yielding a requirement of 139 CT scans for this study. However, only 100 CT scans were available from the hospital database, consisting of 71 males and 29 females. Despite falling short of the calculated ideal, these 100 scans were deemed sufficient and were thus used for the assessment in this study. Sample size was calculated using the standard formula as follows:-

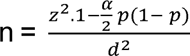 (Lwanga and Lemeshaw 1991)

Where;

n = minimum sample size

z = standard normal deviation, which is 1· 96 at 95% confidence level

p = proportion in the target population, given as 10%

d = sampling error, given as 0.05

By substituting these values into the formula, we get:-

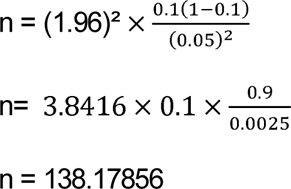

Therefore, the minimum sample size needed for the study, when rounded up is 139.

### Data collection and image processing

Socio-demographic data including age and gender were meticulously gathered for each subject. The measurements of the foramen magnum and occipital condyles were then recorded by a qualified radiographer, ensuring data accuracy. These measurements were obtained from high-resolution, multi-slice cranial CT scans. A total of 100 scans were analyzed, providing ten distinct dimensions of the foramen magnum and occipital condyles for evaluation. For data analysis and visualization, the Digital Imaging and Communications in Medicine (DICOM) images were transferred to a CD plate and subsequently reviewed using a dedicated image viewer.

### Cranial Computed Tomography (CT) imaging protocol

All subjects underwent assessment using a multi-slice NeuViz 16 scanner (Neusoft, N16E130043). During the scanning procedure, subjects were positioned supine to ensure optimal image quality. The images obtained were visually examined to note variations in the shapes of the foramen magnum and occipital condyles. Precise measurements were captured using the same NeuViz 16 multi-slice CT scanner, with a set scanning parameter of 120 kilovolt peak (kVp) and 40 milliamperes-seconds (mAs).

### Foramen magnum and occipital condyles measurements

The foramen magnum length was defined as the greatest internal length along the mid-sagittal plane (Figure 2A, A1-A2). Correspondingly, the foramen magnum width was determined as the widest internal measurement along the transverse plane (Figure 2A, B1-B2). The area of the foramen magnum was calculated using Radinsky’s formula: ¼ × π × Width × Length, while the circumference was estimated by tracing along the bony border of the foramen magnum (Figure 2D).

Moreover, the right occipital condyle’s length was ascertained as the longest measurement along the articular surface, perpendicular to its width (Figure 2B, D1-D2). Its width was measured along the articular surface, perpendicular to the length (Figure 2B, D3-D4). For the left occipital condyle, the length was captured as the greatest measurement along the articular surface, perpendicular to the width (Figure 2B, C1-C2). Meanwhile, the width was quantified along the articular surface, perpendicular to the length (Figure 2B, C3-C4).

Additionally, the minimum intercondylar distance, or the shortest distance between the medial edges of the articular surfaces of the occipital condyles perpendicular to the mid-sagittal plane, was recorded (Figure 2C, E3-E4). The maximum intercondylar distance, which was defined as the longest distance between the medial margins of the occipital condyles perpendicular to the mid-sagittal plane, was also measured (Figure 2C, E1-E2).

**Figure 1:**
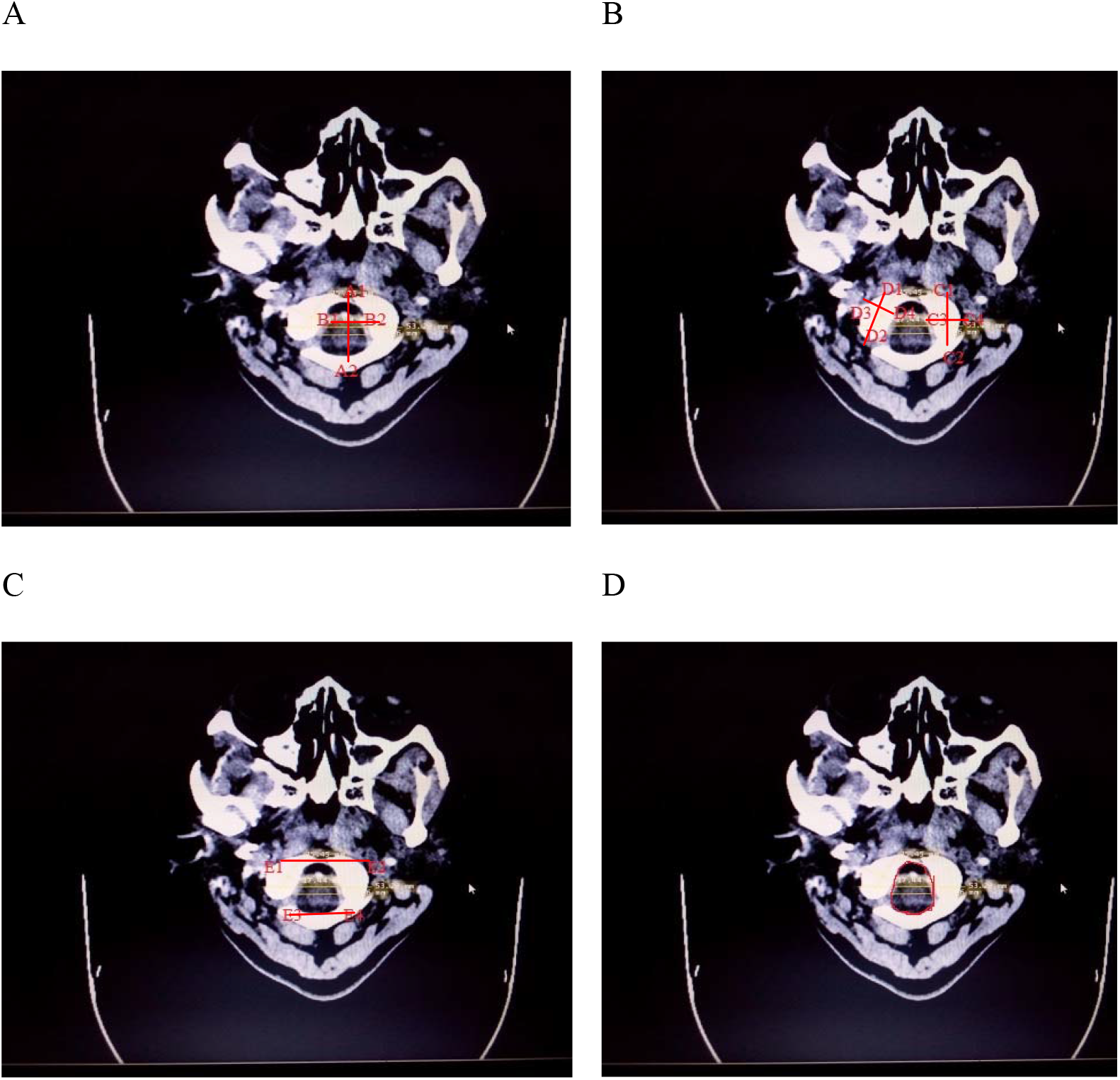
**Illustration of the measured parameters on computed tomographic images**. (A) Foramen magnum length (A1-A2) and width (B1-B2). (B) Left occipital condyle length (C1-C2) and width (C3-C4), and right occipital condyle length (D1-D2) and width (D3-D4). (C) Maximum (E1-E2) and minimum (E3-E4) intercondylar distances. (D) Depicts the area and circumference of the foramen magnum.

### Image analysis and statistical approach

Images generated were meticulously analysed employing DICOM image viewer software. All statistical computations were conducted utilizing the Statistical Package for Social Sciences (SPSS) version 21. An initial normality test was performed on the morphometric measurements for the Foramen Magnum (FM) and Occipital Condyle (OC) parameters. Differences between genders were assessed using the Mann-Whitney U test. The Chi-square test was employed to evaluate the association between the shapes of the studied variables and distinct ethnic groups. Discriminant function analysis was utilized to predict gender and ascertain the accuracy of gender identification. A Spearman correlation was used to evaluate the relationship between parameters such as the Length of the Foramen Magnum (LFM), Width of the Foramen Magnum (WFM), Length of the Right Condyle (LRC), Length of the Left Condyle (LLC), Width of the Right Condyle (WRC), Width of the Left Condyle (WLC), Maximum Intercondylar distance (MIC), and Minimum Intercondylar distance (MnIC). Demographic data of the subjects were described using mean and standard deviation. A p-value of less than 0.05 was considered to indicate statistical significance.

## Results

### Demographic and morphometric characteristics of study subjects

Table 1 summarizes the sociodemographic profiles and the morphometric dimensions of the foramen magnum (FM) and occipital condyle (OC) among patients at the Bauchi State Specialist Hospital, Bauchi, Nigeria. The study sample comprised of a significantly larger proportion of males (70%) compared to females (30%). The mean age for males was 40.00 ± 11.53 years, while for females it was 33.78 ± 9.79 years, within an age range of 21 to 65 years.

**Table 1:** Descriptive statistics of demographic data and morphometric dimensions of foramen magnum and occipital condyle stratified by gender (n= 100)

| Variable | Gender | Mean ± SD | Min | Max |
| --- | --- | --- | --- | --- |
| Age | Male | $40.00 \pm 11.53$ | 21 | 65 |
| (Years) | Female | $33.78 \pm 9.79$ | | |
| LFM | Male | $26.33 \pm 6.51$ | 13.87 | 46.76 |
| (mm) | Female | $21.58 \pm 4.61$ | 11.68 | 35.60 |
| WFM | Male | $21.05 \pm 5.77$ | 7.80 | 35.05 |
| (mm) | Female | $18.31 \pm 4.36$ | 11.19 | 27.29 |
| AFM | Male | $427.34 \pm 208.85$ | 84.93 | 1286.57 |
| (mm) | Female | $311.65 \pm 124.58$ | 168.04 | 762.65 |
| CFM | Male | $68.22 \pm 14.44$ | 34.00 | 99.79 |
| (mm) | Female | $61.54 \pm 10.35$ | 39.91 | 85.87 |
| LRC | Male | $16.77 \pm 8.83$ | 7.09 | 74.59 |
| (mm) | Female | $14.78 \pm 5.07$ | 7.55 | 31.04 |
| WRC | Male | $7.76 \pm 3.03$ | 3.31 | 16.01 |
| (mm) | Female | $7.98 \pm 3.43$ | 3.38 | 14.86 |
| LLC | Male | $18.28 \pm 5.42$ | 7.65 | 30.16 |
| (mm) | Female | $16.95 \pm 4.77$ | 8.50 | 28.81 |
| WLC | Male | 8.57±3.45 | 3.57 | 16.65 |
| (mm) | Female | 8.57±3.57 | 3.72 | 15.02 |
| MIC | Male | 44.89±7.30 | 28.17 | 59.74 |
| (mm) | Female | 40.79±6.03 | 32.79 | 59.90 |
| MnIC | Male | 22.70±4.89 | 11.28 | 32.90 |
| (mm) | Female | 21.34±5.22 | 13.42 | 33.35 |

SD; standard deviation, Min; Minimum, Max; Maximum

The average measurements for FM and OC dimensions differed between genders across all variables. Specifically, for males, the mean length of the foramen magnum (LFM) was 26.33±6.51 mm, width of the foramen magnum (WFM) was 21.05±5.77 mm, area of the foramen magnum (AFM) was 427.34±208.85 mm, circumference of the foramen magnum (CFM) was 68.22±14.44 mm, length of the right condyle (LRC) was 16.77±8.83 mm, width of the right condyle (WRC) was 7.76±3.03 mm, length of the left condyle (LLC) was 18.28±5.42 mm, width of the left condyle (WLC) was 8.57±3.45 mm, maximum intercondylar distance (MIC) was 44.89±7.30 mm, and the minimum intercondylar distance (MnIC) was 22.70±4.89 mm.

For females, the mean LFM was 21.58±4.61 mm, WFM was 18.31±4.36 mm, AFM was 311.65±124.58 mm, CFM was 61.54±10.35 mm, LRC was 14.78±5.07 mm, WRC was 7.98±3.43 mm, LLC was 16.95±4.77 mm, WLC was 8.57±3.57 mm, MIC was 40.79±6.03 mm, and MnIC was 21.34±5.22 mm. These findings provide a comprehensive overview of the FM and OC dimensions among the male and female patients in this study group. The standard deviations, minimum and maximum values for each dimension are presented for both genders in Table 1.

### Frequency distribution of foramen magnum (FM) and occipital condyle (OC)

The morphological variations of the foramen magnum (FM) and occipital condyle (OC) shapes are depicted in Tables 2 and 3 respectively. In our cohort, the most prevalent shape of the foramen magnum was found to be oval, constituting 33.3% of the sample. This was followed by round (18.2%), egg (12.1%), tetragonal (10.1%), irregular and pentagonal (each 9.1%), and hexagonal (7.1%) shapes.

**Table 2:** Frequency distribution of foramen magnum morphological variations.

| Types | Number | Percentage |
| --- | --- | --- |
| Oval | 33 | 33.3 |
| Round | 18 | 18.2 |
| Egg | 12 | 12.1 |
| Tetragonal | 10 | 10.1 |
| Pentagonal | 9 | 9.1 |
| Hexagonal | 7 | 7.1 |
| Irregular | 10 | 10.1 |
| Total | 99 | 100 |

**Table 3:** Frequency distribution of occipital condyles morphological variations.

| Types | Number | Percentage |
| --- | --- | --- |
| Kidney | 39 | 39 |
| Oval | 14 | 14 |
| 8-like | 10 | 10 |
| S-like | 17 | 17 |
| Ring-like | 10 | 10 |
| Triangular | 10 | 10 |
| Total | 100 | 100 |

For the occipital condyles, the dominant morphological variation was a kidney-like structure, accounting for 39% of the sample. The other shapes observed were the S-like (17%), oval (14%), and 8-like, ring-like, and triangular types (each 10%).

### Variations of foramen magnum and occipital condyles dimensions among gender

Table 4 present the gender-based variations in foramen magnum and occipital condyle dimensions. The Mann-Whitney U test was employed to assess these differences. A statistically significant disparity was observed in the length of the foramen magnum (LFM), width of the foramen magnum (WFM), area of the foramen magnum (AFM), circumference of the foramen magnum (CFM), and maximum intercondylar distance (MIC), with males demonstrating larger dimensions than females (p<0.05).

**Table 4:**
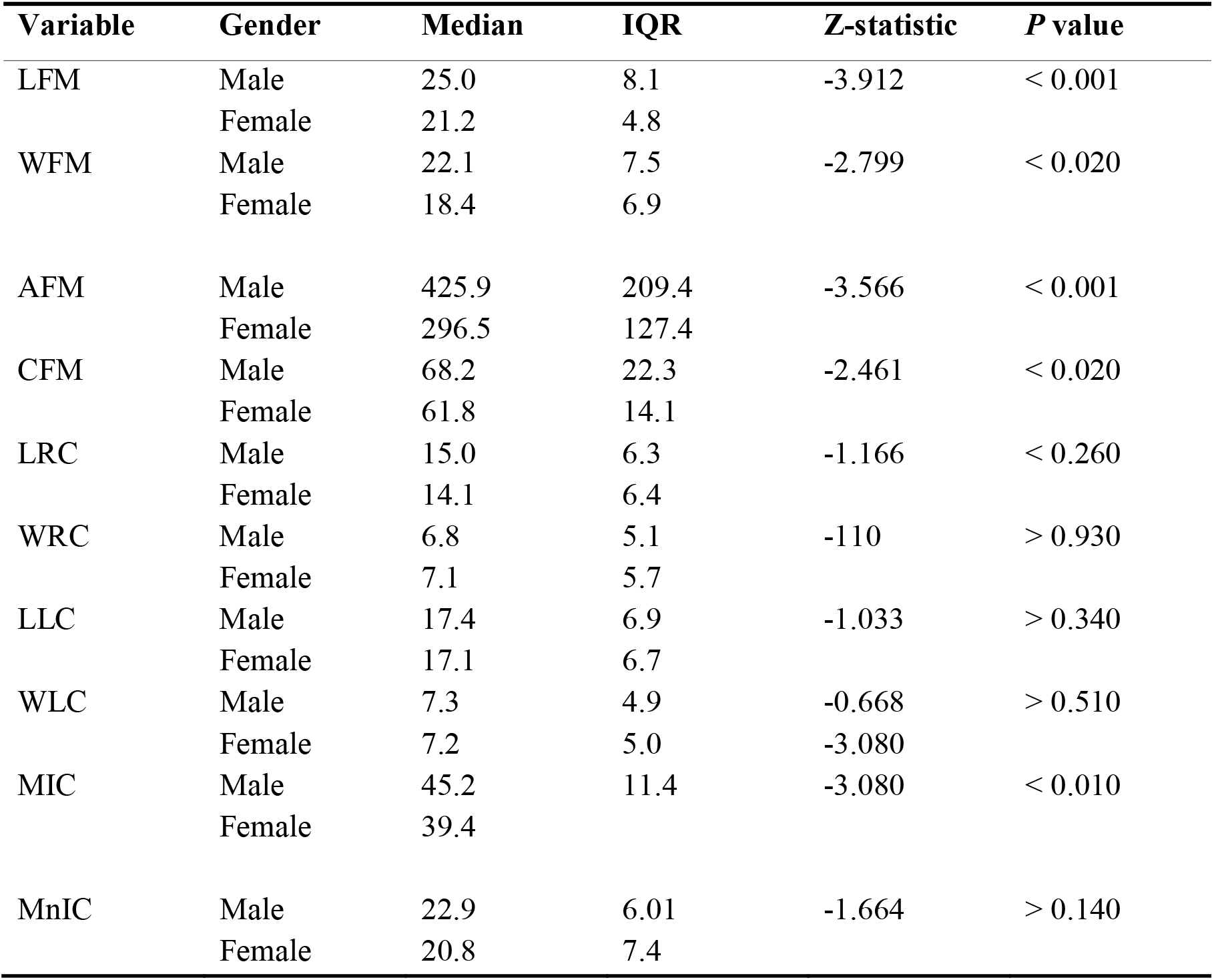
Gender-based variations of foramen magnum and occipital condyles dimensions.

In contrast, no significant difference between genders was observed in the length of right/left occipital condyle (LRC, LLC), width of right/left occipital condyle (WRC, WLC), and minimum intercondylar distance (MnIC) (p>0.05).

### Ethnic variation in the shapes of the foramen magnum

Table 5 presents the association of different shapes of the foramen magnum with various ethnic groups. The distribution of foramen magnum shapes across the ethnic groups was evaluated using the chi-square (X2) test. The test produced a chi-square statistic of 44.135 with 54 degrees of freedom (df). The p-value was determined to be 0.829, which is above the commonly used significance level of 0.05, suggesting that the shapes of the foramen magnum do not significantly differ across the ethnic groups investigated in this study.

**Table 5:**
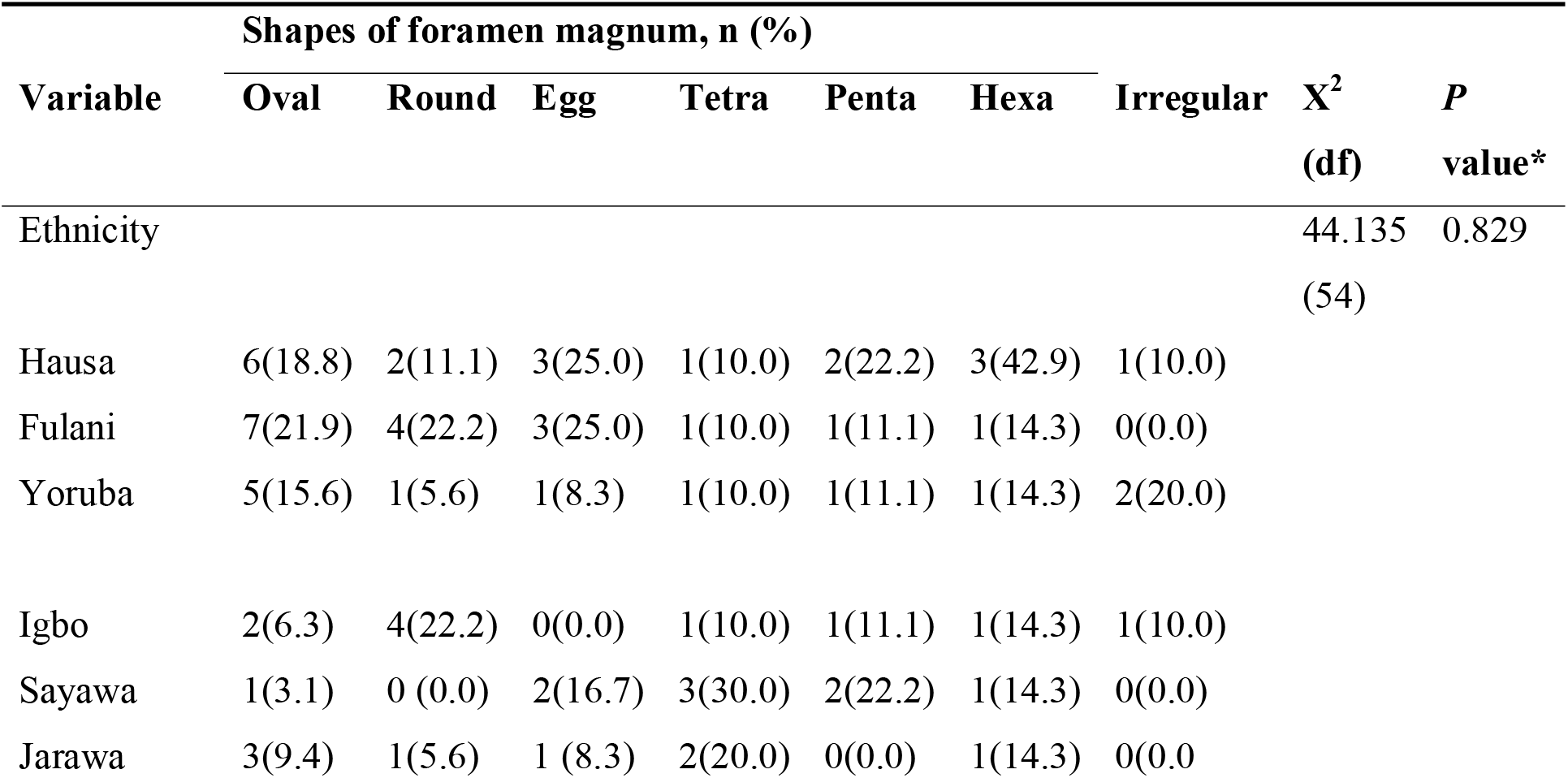
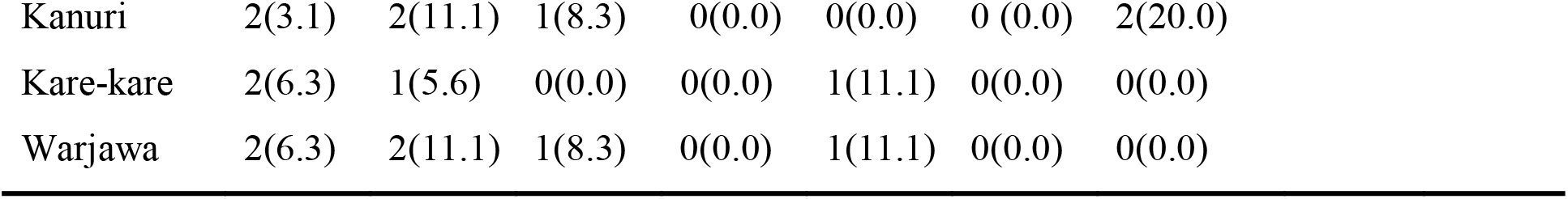
Association of foramen magnum shapes and ethnicity.

| Variable | Shapes of foramen magnum, n (%) |  |  |  |  |  |  | X<sup>2</sup><br>(df) | P<br>value* |
| --- | --- | --- | --- | --- | --- | --- | --- | --- | --- |
|  | Oval | Round | Egg | Tetra | Penta | Hexa | Irregular |  |  |
| Ethnicity |  |  |  |  |  |  |  | 44.135<br>(54) | 0.829 |
| Hausa | 6(18.8) | 2(11.1) | 3(25.0) | 1(10.0) | 2(22.2) | 3(42.9) | 1(10.0) |  |  |
| Fulani | 7(21.9) | 4(22.2) | 3(25.0) | 1(10.0) | 1(11.1) | 1(14.3) | 0(0.0) |  |  |
| Yoruba | 5(15.6) | 1(5.6) | 1(8.3) | 1(10.0) | 1(11.1) | 1(14.3) | 2(20.0) |  |  |
| Igbo | 2(6.3) | 4(22.2) | 0(0.0) | 1(10.0) | 1(11.1) | 1(14.3) | 1(10.0) |  |  |
| Sayawa | 1(3.1) | 0 (0.0) | 2(16.7) | 3(30.0) | 2(22.2) | 1(14.3) | 0(0.0) |  |  |
| Jarawa | 3(9.4) | 1(5.6) | 1 (8.3) | 2(20.0) | 0(0.0) | 1(14.3) | 0(0.0) |  |  |

|  |  |  |  |  |  |  |  |
| --- | --- | --- | --- | --- | --- | --- | --- |
| Kanuri | 2(3.1) | 2(11.1) | 1(8.3) | 0(0.0) | 0(0.0) | 0 (0.0) | 2(20.0) |
| Kare-kare | 2(6.3) | 1(5.6) | 0(0.0) | 0(0.0) | 1(11.1) | 0(0.0) | 0(0.0) |
| Warjawa | 2(6.3) | 2(11.1) | 1(8.3) | 0(0.0) | 1(11.1) | 0(0.0) | 0(0.0) |

It can be observed that the oval shape of the foramen magnum is most common among the Fulani ethnic group, while the Hexagonal shape is most common in the Hausa group. The Igbo ethnic group showed a higher frequency of the round shape, while the Sayawa group showed a higher frequency of the Tetragonal shape. However, it’s important to note that due to the p-value being above 0.05, these apparent trends do not represent a statistically significant association between ethnicity and the shape of the foramen magnum. Therefore, these observations could have resulted from random variation within the sample population. Further investigation with a larger sample size may be needed to determine whether there are meaningful differences in foramen magnum shape across different ethnic groups.

### Ethnic variation in the shapes of the occipital condyles

Table 6 investigates the association between different shapes of occipital condyles and various ethnic groups. The distribution of occipital condyle shapes across the ethnic groups was evaluated using the chi-square (X2) test, which yielded a statistic of 42.112 with 45 degrees of freedom (df). The resulting p-value was 0.595, exceeding the conventional significance level of 0.05, indicating that the occipital condyle shapes do not significantly differ among the ethnic groups examined in this study.

**Table 6:**
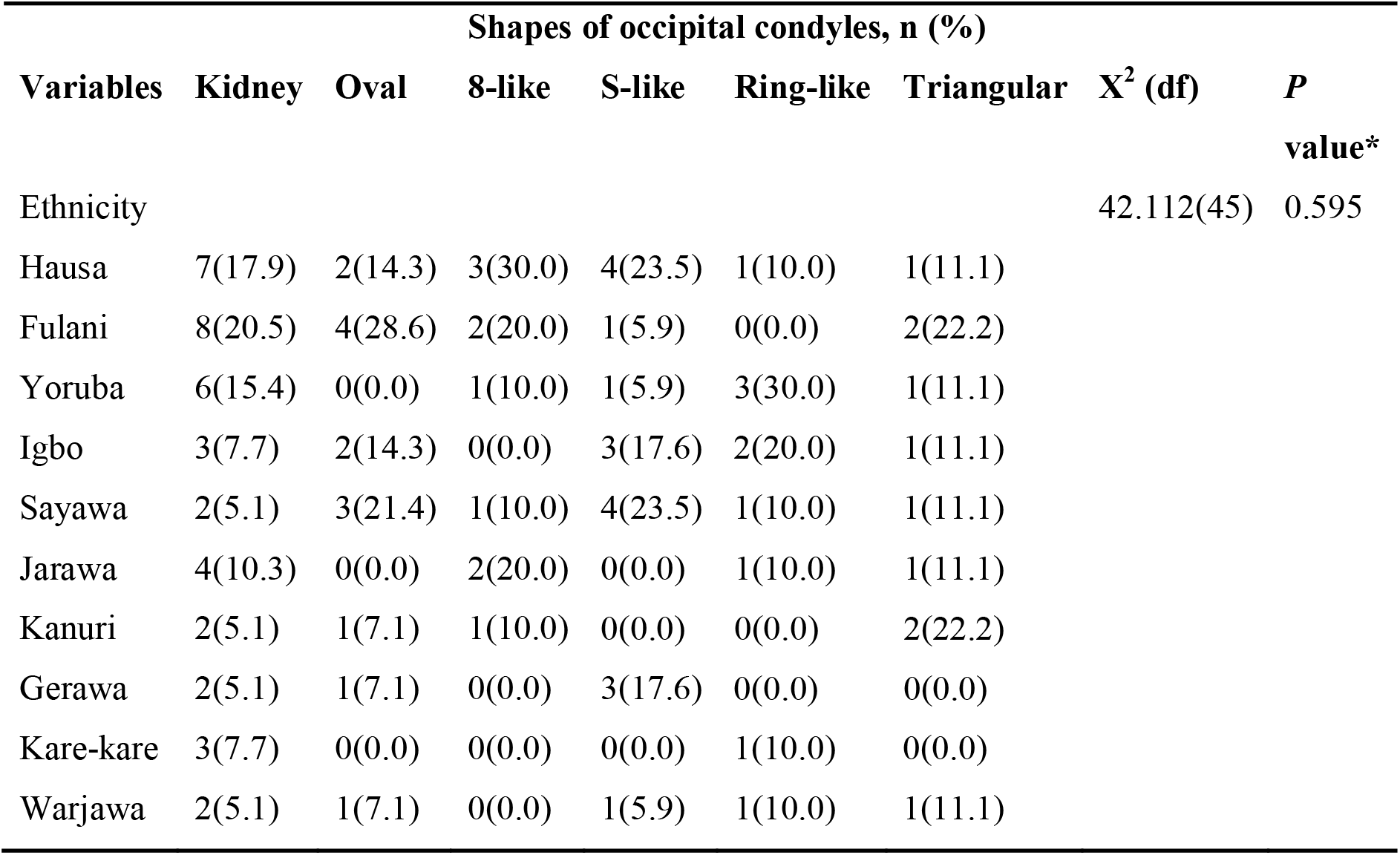
Association of occipital condyles shapes and ethnicity.

| Variables | Shapes of occipital condyles, n (%) |  |  |  |  |  | X<sup>2</sup> (df) | P value* |
| --- | --- | --- | --- | --- | --- | --- | --- | --- |
|  | Kidney | Oval | 8-like | S-like | Ring-like | Triangular |  |  |
| Ethnicity |  |  |  |  |  |  | 42.112(45) | 0.595 |
| Hausa | 7(17.9) | 2(14.3) | 3(30.0) | 4(23.5) | 1(10.0) | 1(11.1) |  |  |
| Fulani | 8(20.5) | 4(28.6) | 2(20.0) | 1(5.9) | 0(0.0) | 2(22.2) |  |  |
| Yoruba | 6(15.4) | 0(0.0) | 1(10.0) | 1(5.9) | 3(30.0) | 1(11.1) |  |  |
| Igbo | 3(7.7) | 2(14.3) | 0(0.0) | 3(17.6) | 2(20.0) | 1(11.1) |  |  |
| Sayawa | 2(5.1) | 3(21.4) | 1(10.0) | 4(23.5) | 1(10.0) | 1(11.1) |  |  |
| Jarawa | 4(10.3) | 0(0.0) | 2(20.0) | 0(0.0) | 1(10.0) | 1(11.1) |  |  |
| Kanuri | 2(5.1) | 1(7.1) | 1(10.0) | 0(0.0) | 0(0.0) | 2(22.2) |  |  |
| Gerawa | 2(5.1) | 1(7.1) | 0(0.0) | 3(17.6) | 0(0.0) | 0(0.0) |  |  |
| Kare-kare | 3(7.7) | 0(0.0) | 0(0.0) | 0(0.0) | 1(10.0) | 0(0.0) |  |  |
| Warjawa | 2(5.1) | 1(7.1) | 0(0.0) | 1(5.9) | 1(10.0) | 1(11.1) |  |  |

In the observed data, the kidney shape of the occipital condyles was most frequently seen in the Fulani ethnic group, while the 8-like shape was most common among the Hausa group. The Igbo and Sayawa ethnic groups showed a higher frequency of the S-like shape, whereas the ring-like shape was more frequent in the Yoruba group. The triangular shape was equally distributed among Hausa, Fulani, Yoruba, Igbo, Sayawa, Jarawa, and Warjawa.

However, the above p-value signifies these trends do not denote a statistically significant association between ethnicity and the shape of occipital condyles. As such, these observations could potentially be the product of random variation within the sample population. A more comprehensive investigation with a larger sample size might be required to determine if there are significant differences in occipital condyle shapes across various ethnic groups.

### Gender discrimination through discriminant function analysis

The results in Tables 7 and 8 provide insight into the gender differentiation capabilities of the foramen magnum and occipital condyle dimensions based on computed tomography (CT) scans.

**Table 7:** Discriminant function analysis using foramen magnum and occipital condyle dimensions in CT scan.

| Variable | Male | | Female | | Wilki's $\lambda$ | F | P value* |
| --- | --- | --- | --- | --- | --- | --- | --- |
|  | Mean | SD | Mean | SD |  |  |  |
| LFM | 26.35 | 6.47 | 21.45 | 4.50 | 0.876 | 13.875 | 0.001 |
| WFM | 21.19 | 5.87 | 17.74 | 4.72 | 0.925 | 7.970 | 0.006 |
| AFM | 430.99 | 209.63 | 300.66 | 126.89 | 0.910 | 9.720 | 0.002 |
| CFM | 68.37 | 14.49 | 61.36 | 10.32 | 0.946 | 5.611 | 0.020 |
| LRC | 16.72 | 8.78 | 14.51 | 5.01 | 0.984 | 1.610 | 0.208 |
| WRC | 7.72 | 3.03 | 7.77 | 3.42 | 1.000 | 0.003 | 0.953 |
| LLC | 18.24 | 5.39 | 16.74 | 4.66 | 0.983 | 1.710 | 0.184 |
| WLC | 8.54 | 3.44 | 8.29 | 3.60 | 0.999 | 0.103 | 0.749 |
| MIC | 44.90 | 7.25 | 40.26 | 6.15 | 0.914 | 9.190 | 0.003 |
| MnIC | 22.72 | 4.86 | 20.90 | 5.30 | 0.973 | 2.751 | 0.100 |

**Table 8:**
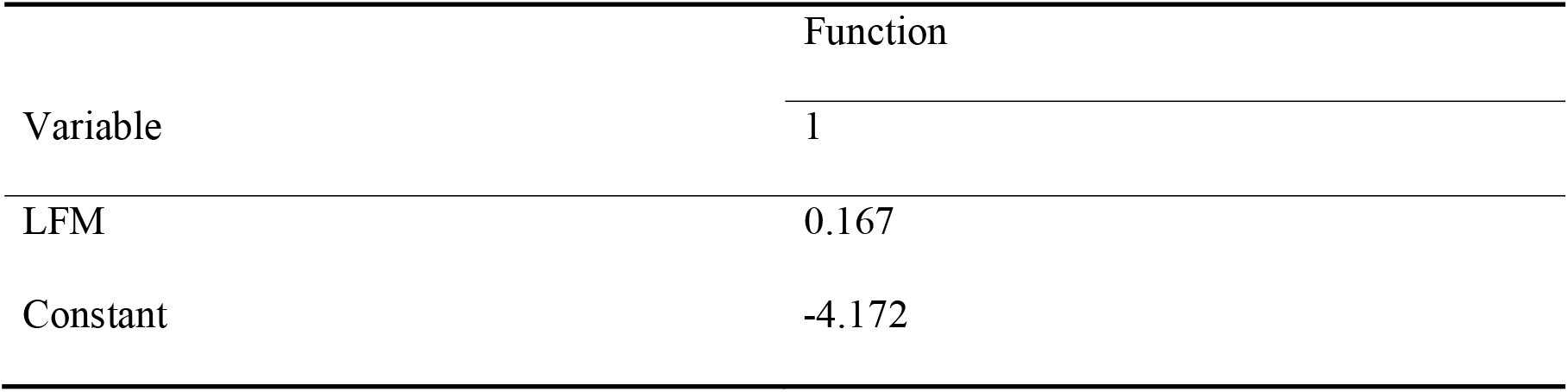
Canonical discriminant function coefficient.

| Variable | Function |
| --- | --- |
|  | 1 |
| LFM | 0.167 |
| Constant | -4.172 |

Table 7 compares the mean and standard deviation (SD) values of these dimensions between male and female participants. The gender discrimination significance for each variable is assessed using the Wilk’s lambda test, F-value, and the corresponding p-value. The lower the Wilk’s lambda, the more significant the variable is in distinguishing the two groups. The F-value assesses the degree of variation between groups compared to the variation within groups. The length of the foramen magnum (LFM) had the greatest contribution to gender discrimination, as evidenced by the smallest Wilk’s lambda value (0.876), highest F-value (13.875), and a p-value less than 0.001. This shows that there is a statistically significant difference between males and females concerning the LFM, and it effectively discriminates between the two genders.

In contrast, variables such as WRC, WLC, and MnIC exhibit high Wilk’s lambda values and non-significant p-values, indicating they do not significantly contribute to gender differentiation in this context.

### Efficacy of foramen magnum length in predicting gender

Table 9 presents the results of the predictive accuracy of using the foramen magnum length to determine gender within the Bauchi population. The results are divided into predicted group membership versus actual gender, with values indicating the number of correctly sexed cases Out of 71 male cranial CT scans, 45 were correctly identified as male, resulting in an accuracy rate of 63.4%. However, 26 male cases were incorrectly classified as female, indicating a misclassification rate of 36.6%.

**Table 9:**
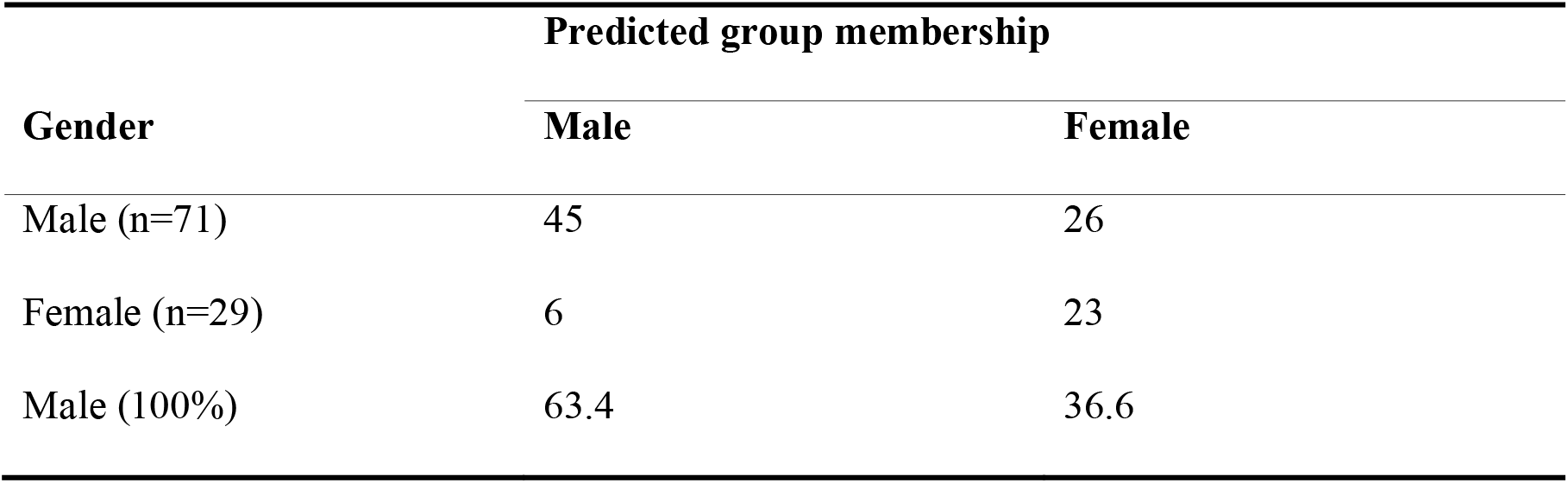
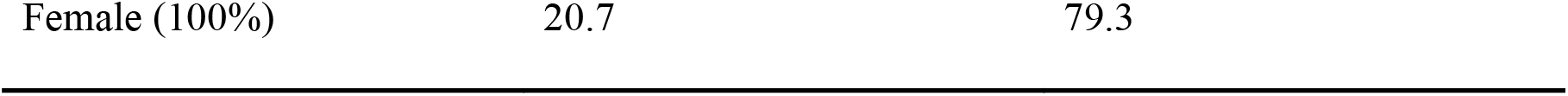
Accuracy of foramen magnum length for gender prediction in Bauchi population.

On the other hand, the predictive accuracy was higher for females. Out of 29 female cranial CT scans, 23 were accurately identified as female, giving an accuracy rate of 79.3%. However, 6 female cases were incorrectly classified as male, denoting a misclassification rate of 20.7%. While the foramen magnum length was a significant predictor of gender, as demonstrated in previous tables, the accuracy of gender identification varied between males and females. The accuracy was higher for females compared to males in the studied population. This suggests that the discriminant function using the foramen magnum length is more reliable for sexing female crania than male in this population. However, overall, it correctly classified 68.0% of all cases, demonstrating its utility in gender identification in the Bauchi population.

### Correlation analysis of foramen magnum and occipital Condyle Dimensions

Table 10 explores the correlations between different dimensions of the foramen magnum and occipital condyles. There is a statistically significant strong correlation between Length of Foramen Magnum (LFM) and Length of Right Condyle (LRC) (r=0.279, p<0.005), suggesting that as the length of the foramen magnum increases, the length of the right condyle also increases in a meaningful way.

**Table 10:** Relationship of foramen magnum and occipital condyle dimensions.

| **Variable** | **correlations** | **LRC** | **WRC** | **LLC** | **WLC** | **MIC** | **MniC** |
| --- | --- | --- | --- | --- | --- | --- | --- |
| LFM | r | 0.279 | 0.194 | 0.252 | 0.231 | 0.316 | 0.220 |
|  | p value | 0.001 | 0.053 | 0.012 | 0.021 | 0.001 | 0.028 |
| WFM | r | 0.337 | 0.203 | 0.296 | 0.264 | 0.404 | 0.259 |
|  | p value | 0.001 | 0.042 | 0.003 | 0.008 | 0.001 | 0.009 |

Moreover, a similar trend can be observed with LFM and MIC (r=0.316, p<0.001), suggesting a significant correlation between these two measurements. The Width of Foramen Magnum (WFM) also shows strong correlations with several other dimensions. Notably, there is a strong relationship between WFM and LRC (r=0.337, p<0.001), Width of Left Condyle (WLC) (r=0.264, p<0.008), and MIC (r=0.404, p<0.001).

Thus the results revealed significant relationships between various foramen magnum and occipital condyle dimensions, providing key insights into the intricate anatomical relationships between these structures in among Bauchi population.

## Discussion

This comprehensive study provides a detailed examination of the foramen magnum and occipital condyle dimensions within the Bauchi population, encompassing an array of morphological variations and their potential significance in sex determination.

As our study demonstrates, the foramen magnum’s most prevalent shapes were oval and round, mirroring the results of previous research (Garcia *et al.,* 2011). Similarly, the occipital condyle shapes noted in our sample - kidney, oval, 8-like, S-like, ring-like, and triangular -align with the shapes identified in other studies (Kumar *et al*., 2019) (Ozer et al., 2011), though there was a discrepancy in the most common shape, with our study identifying the kidney shape as the most common, in contrast to the oval-like shape identified by Ozer et al.

Sex-related differences in foramen magnum dimensions were evident in our research. We discovered that the mean Length and Width of the Foramen Magnum (LFM, WFM) were larger in males than in females, echoing the results of with (El-Barrany *et al*., 2016). However, when compared to previous studies (Gopalro *et al*., 2013, Uthman *et al*., 2014 and El-Barrany *et al*., 2016), the dimensions found in our study were lower. This disparity could be attributed to differences in population, methodology, or measurement techniques and further emphasizes the need for population-specific studies.

Our findings also reveal that the mean Area of Foramen Magnum (AFM) of male skulls (427.34± 208.85mm²) is significantly larger than female skulls (311.65±124.58mm²) (Table 1 and 4). Even though the AFM values in our study were lower than those reported in previous studies (Gunay *et al*., 2000) in both male and female (909.9 and 819.0mm²), (Catalina and Herrera 1987) (888.4 and 801.0mm²), (Routal *et al*., 2014) (819.0 and 771.0mm²) and (Salih *et al*., 2015) (819.84 and 692.13mm²), the trend of males having larger AFM than females remains consistent.

Interestingly, we found that the width of the occipital condyles (CFM) was broader in females than in males (68.22±14.44 and 61.54 ±10.35) (Table 1), a finding that contradicts earlier studies but aligns with (Sneha *et al.,* 2014). The variability in these results underlines the complexity of the biological factors shaping these structures and calls for further research to elucidate these sex-based discrepancies.

In terms of sex determination, our study indicates that LFM could predict gender in the Bauchi population with an overall accuracy rate of 68.0%. This finding corroborates other research that identified single variables as significant gender discriminators (Singh and Talwar, 2013) (Macaluso, 2010) (Wani *et al*., 2021). Moreover, our study establishes significant correlations between various foramen magnum and occipital condyle dimensions, lending further credence to the hypothesis that these structures could be used for sex determination. However, some relationships such as the correlation between the LFM and LLC or the width of the right and left occipital condyle were not found to be significant, emphasizing the need for continued research in this field.

In conclusion, our findings contribute to the growing body of literature emphasizing the value of foramen magnum and occipital condyle morphology and dimensions in anthropological and forensic contexts. Additionally, variations in dimensions should be taken into consideration when performing clinical and radiological diagnoses and during a surgical procedure. This study also reveals that the shape of the foramen magnum and the occipital condyle is not a mark of a specific ethnic group. Future studies should consider these findings when examining morphological sex differences and explore the potential of these structures as markers of sexual dimorphism.

## Authors contribution

**Fatima Muhammad**: Data curation, Writing - original draft. **Umar Ahmad**: Conceptualisation, Methodology, Resources, Writing- Review & Editing; Supervision. **Tawfiq Yousef Tawfiq ZYOUD:** Methodology, Writing- Review & Editing. **Murtala Muhammad Jibril**: Writing- Review & Editing. **Suleiman Hamidu Kwairanga**: Writing- Review & Editing.

## Data Availability

All data produced in the present work are contained in the manuscript

